# Effects of Singing on Vascular Health in Older Adults with Coronary Artery Disease: A Randomized Trial

**DOI:** 10.1101/2024.07.25.24311033

**Authors:** Mehri Bagherimohamadipour, Muhammad Hammad, Alexis Visotcky, Rodney Sparapani, Jacquelyn Kulinski

**Affiliations:** Division of Cardiovascular Medicine, Medical College of Wisconsin, Milwaukee, WI; Department of Biostatistics, Medical College of Wisconsin, Milwaukee, WI

**Keywords:** singing, cardiac rehabilitation, endothelial function, vascular function, heart rate variability

## Abstract

**Background:** The impact of singing on cardiovascular health has not been extensively studied. The aim of this study is to investigate the effect of singing on cardiovascular biomarkers in an aging population with coronary artery disease.

**Methods:** Participants had three study visits separated by 2-7 days, according to a randomized, single-blind, cross-over, controlled design: (1) a 30-minute period of coached singing from an in-person music therapist, (2) a 30-minute period of singing along to an instructional video and (3) a 30-minute rest (control). Primary outcomes included macrovascular endothelial function assessed by brachial artery flow-mediated dilation and microvascular function assessed by peripheral arterial tonometry (Framingham reactive hyperemia index; fRHI). Heart rate variability was a secondary outcome.

**Results:** Sixty-five subjects (mean age 67.7± 0.8, 40% women) completed the study. Compared to control, there was an increase in fRHI for the singing video intervention (estimate 0.54, SE 0.25, p=0.005) but not for the coaching intervention (estimate 0.11, SE 0.18, p=0.570). There was no change in macrovascular function with either intervention. The low frequency/high frequency (LF/HF) ratio increased by 2.80 (SE 1.03, p=0.008), and the LnHF power decreased by −0.90 ms^2^ (SE 0.29, p=0.003) with the video (during to pre-change). When assessing post- to pre- change, the coaching intervention showed a significant change of −0.62 ms^2^ (SE 0.29, p=0.036) in LnHF power.

**Conclusions:** Singing along to an instructional video for 30 minutes improved microvascular, but not macrovascular, endothelial function, in older patients with CAD. HRV changes with singing are similar to that of exercise.

## Introduction

Exercise promotes healthy vascular aging and is a crucial means for prevention, treatment, and rehabilitation of cardiovascular disease (CVD). According to data from the 2020 National Health Interview Survey in the U.S., fewer than one-fourth (24.2%) of adults meet national recommendations for physical activity (at least 150 minutes of moderate-intensity exercise per week).^1^ However, this percentage is markedly lower for adults ages 65 and over, with only 15.3% of men and just 10.8% of women meeting national recommendations for physical activity.^1^ Similarly, older adults are less likely to participate in cardiac rehabilitation (CR) programs, even when referred.^2–6^ CVD in older adults is usually complicated by age-related complexities, including multi-morbidity, polypharmacy, frailty, deconditioning, falls, disability, and other challenges that make participation (or the perception of participation) in CR difficult.^4,7^ Alternative forms of therapy to reduce CVD burden and improve health are needed in this aging population.

Music as a therapeutic is attractive for a variety of reasons including minimal risk to patients, ease of use, accessibility, and pervasiveness across cultures. The physiological demands of singing are comparable with walking at a moderately brisk pace, suggesting that the health benefits with singing may overlap with that of exercise.^8^ Unlike traditional physical exercise, the impact of singing on cardiovascular health has not been extensively studied. Vascular endothelial function strongly predicts cardiovascular events in patients with and without CVD,^9^ and endothelial dysfunction can be reversed by interventions (such as physical exercise) known to reduce CVD risk.^10,11^ Our team has been the first to demonstrate that just 14 minutes of solo singing improves endothelial function acutely, regardless of singing expertise.^12^ Changes were more robust in subjects with baseline endothelial dysfunction. Major limitations of the prior study included lack of randomization and a control arm.

HRV is the variability between R-R intervals in successive heartbeats and is the result of a complex interaction between respiratory activity and autonomic cardiovascular control between the two branches of the autonomic nervous system (sympathetic and parasympathetic).^13^ Measurements of HRV have been found to be powerful predictors of cardiac morbidity and mortality.^14,15^ While singing has been shown to improve HRV in young, healthy populations,^16^ there is no data on the impact of singing on HRV in patients with CVD. This is a secondary outcome measure that we plan to explore here.

Live music engages listeners to a greater extent than pre-recorded music.^17^ It is unknown whether live music has different physiological effects compared to pre-recorded music. Because we hypothesize that some of the benefits of physical exercise may extend to the physical activity of singing, incorporating a singing coach (to increase level of exertion) may be more impactful.

The aim of this study is to investigate the impact of singing on important cardiovascular biomarkers, including vascular endothelial function and heart rate variability, in an aging population with established coronary artery disease (CAD). The study consists of three arms, according to a randomized, single-blind, crossover, controlled design. Participants each had three visits on three different occasions for the following interventions: (1) a 30-minute period of coached singing from an in-person, board-certified music therapist, (2) a 30-minute period of singing along to an instructional video and (3) a 30-minute rest (control) visit.

## Methods

### Subject Recruitment

All subject-related study activities were performed at a large academic medical center after approval by the local Institutional Review Board (IRB). Subjects were identified by reviewing the electronic medical records of patients visiting the outpatient cardiology clinics. To increase enrollment of women, as well as black, and Hispanic patients, of both genders, potential subjects were also identified using a cohort discovery tool (TriNetX) from the institution’s Clinical Research Data Warehouse. The Honest Broker tool was used to extract desired patient information including patient demographics, medication history, diagnosis/problem list, provider notes and lab/imaging results for each identified subject.

The eligibility criteria for the study included patients between 55 and 79 years of age with a history of coronary artery disease defined as having a history of myocardial infarction, coronary artery stenosis greater than 50%, percutaneous coronary intervention with stent placement and/or balloon angioplasty, or coronary artery bypass grafting. We excluded patients who had a permanent pacemaker or implantable cardioverter defibrillator (ICD), history of atrial fibrillation, atrial flutter or atrial tachycardia, Parkinson’s disease or a tremor, amputated upper extremity or presence of upper-arm (dialysis) fistula, fingernail onychomycosis (fungal infections resulting in thickening of the nails), pregnancy, current illicit drug use (marijuana, tobacco, cocaine, amphetamines, etc.), current excessive alcohol use (defined as more than 14 drinks/week for women, more than 28 drinks/week for men), unstable coronary heart disease (active symptoms of chest discomfort), history of a stroke or transient ischemic attack (TIA), peripheral arterial disease, known history of cognitive impairment or inability to follow study procedures, cancer requiring systemic treatment within five years of enrollment, subjects requiring supplemental oxygen use, dosing changes of vasoactive medications in the 6 weeks prior to enrollment, and non-English speaking subjects (instructional videos with lyrics were recorded in English).

### General Study Design

Eligible patients meeting the inclusion and exclusion criteria were contacted via phone or email to provide a brief overview of the study, to assess their interest in the study and to determine initial eligibility in the form of a short screening questionnaire. A detailed screening visit was then completed via a phone call to review the informed consent, the patient’s medical history, and to schedule their three in-person study visits. The medical history form included demographics, medical history, current medications, and physical limitations and/or mobility issues.

At the first study visit, a written informed consent was obtained prior to initiating any study related activities, and the subject underwent randomization in a single-blind, control, crossover design fashion. The participants were randomized, stratified by gender, using an electronic randomization scheme created by an independent statistician not associated with this study. Patients were asked to come to all three study visits fasting from food or beverage (other than water) for 8 hours prior to the visit and were asked to hold their morning medications for the vascular function measurements (all study visits were conducted in the morning). Baseline vital signs (heart rate, blood pressure, pulse oximetry), weight and height, and baseline measurements of heart rate variability (HRV), brachial artery flow-mediated dilation (FMD), and peripheral arterial tonometry (PAT), were taken prior to the intervention.

To ensure rigor of the study, all study personnel including the principal investigator, biostatisticians, sonographers, and the voice professor were kept blinded to the intervention. The only two people unblinded to the intervention were the research coordinator, who scheduled and conducted the study visits, and the music therapist, who led the live singing sessions. The music therapist and research coordinator were not involved in data analysis.

### Singing protocol

The subjects attended two different singing interventions visits and one control visit (rest period), each 30 minutes in duration, in three separate visits according to randomized, controlled, single-blind, crossover design. All interventions were conducted with the subject in a seated position in a private exam room. The visits were separated by a minimum of 2 days (up to 7 days) to allow for potential wash-out effects of prior intervention visits.

A video series was specifically created and recorded for the purposes of this study. The video singing intervention included an instructional sing-along video with a voice professor playing the piano and directing an elderly student in singing. The 10-minute vocal warm-up included semi-occluded vocal tract and breathing exercises. The subjects selected 2 songs to sing for 10 minutes each from four different music genres including Folk (*This Land is Your Land)*, Pop *(Hey Jude)*, Country *(Jolene)*, and Hymn *(Amazing Grace)* - varying in tempo, melodic contour, and rhythm to fill the full 10 minutes per song. The coach intervention included a live in-person singing session with a board-certified music therapist, who alternated between the keyboard or guitar depending on the subjects’ music selection. This session also included the 10-minute vocal warm-up followed by two songs at 10 minutes each. Song choices were selected by the subject from a multi-genre binder targeting adults ages 55-79 and including 40 song choices (Supplementary Table 1). The music therapist encouraged participation and increased exertion during the session. During the control intervention, subjects had a 30-minute period of rest, sitting upright as they would be for the singing intervention. During this rest period, they were not allowed to sleep, watch television, read, browse their smartphone, or listen to music. During this visit, study subjects underwent a validated, tablet-based screening test for hearing loss.^18^

### Measures of Vascular Endothelial Function

Vascular function measurements were performed before and after each singing and rest intervention with subjects in the supine position. The brachial artery flow mediated dilation (FMD) technique^9^ was used to assess macrovascular function, and peripheral arterial tonometry (PAT)^19,20^ was used to assess microvascular function. FMD is a non-invasive measure of vascular endothelial function that uses high-resolution ultrasound technology (7.5-13 Hz probe) to measure changes in brachial artery diameter in response to reactive hyperemia after a 5-minute period of arterial occlusion (>200 mm Hg systolic or at least 60 mm Hg above resting systolic blood pressure with the blood pressure cuff below the elbow). On rapid deflation of the cuff, transient hyperemia stimulates nitric oxide production and release from the endothelium, resulting in dilation. The protocol has been previously described in detail.^21^ FMD is expressed as the change in post-stimulus diameter as a percentage of the baseline diameter (FMD%). Because there is a high degree of technical skill related to assessing FMD, we routinely assess the reproducibility of these measurements in our lab and have shown excellent reproducibility across technicians.^22^

Microvascular endothelial function by digital PAT (Endo-PAT 2000, Itamar Medical, Israel) is expressed as reactive hyperemia index (RHI) and Framingham reactive hyperemia index (fRHI).^19,23^ Endothelium-mediated changes in vascular tone after occlusion of the brachial artery reflect a downstream hyperemic response.^24^ Disposable, pneumatic finger probes were placed on both index fingers. Each recording included 5 minutes of baseline measurement, 5 minutes of occlusion, and 5 minutes post-occlusion (hyperemic period). PAT measurements were taken simultaneously with FMD.

We used a nitroglycerin mediated dilation (NMD) test to differentiate between endothelium-dependent vasodilation and endothelium-independent vasodilation.^25^ Measurement of blood vessel diameter was done before and after the subject was given a pill of nitroglycerin (0.4 mg) sublingually (no blood pressure cuff inflation). NMD was only done once during each study visit, after the second FMD and PAT measurements were completed. NMD was optional and was not given if the participant met at least one of the following criteria: systolic blood pressure less than 100 mmHg; prior intolerance or adverse reaction to nitroglycerin; history of migraine headaches; or sildenafil, tadalafil, or vardenafil use within a week of participating in the study.

### Heart rate variability (HRV)

An appropriately sized (Bluetooth-capable) chest strap (Polar, Kempele, Finland) with a heart rate sensor was applied to the subject’s bare chest. Three-minute-long HRV recordings were obtained before, during (20-25 minutes in), and after singing (or rest control). The data was transmitted to an iPad using the Elite HRV (Asheville, NC). The normal-to-normal (NN) intervals included all intervals between adjacent QRS complexes resulting from sinus node depolarizations. HRV (time domain) was reported as the standard deviation of NN intervals (SDNN) and the root mean square of the successive differences (RMSSD). HRV (frequency domain) was reported as low frequency (LF) power (%), high frequency (HF) power (%), LF/HF ratio, and natural log (HF power).

### Other measurements

Perceived exertion with singing was reported by study subjects using the Borg Rating of Perceived Exertion (RPE) scale.^26^ The Borg RPE is a well-validated, qualitative scale used to assess an individual’s level of exertion and is easy to understand. The scale ranges from 6 to 20, whereas 6 means “no exertion at all” and 20 means “maximal exertion”. At the end of each intervention, subjects were asked to rate their perceived level of exertion using this scale. The Borg RPE is the preferred method to assess intensity among those individuals who take medications that affect heart rate or pulse due to the scale’s ability to capture exertion from central cardiovascular, respiratory, and nervous system functions.^27^ At the end of the singing intervention, subjects were asked to rate their perceived exertion level using the Borg RPE.

### Sample size calculations

The pre-specified primary outcome was percent brachial flow-mediated dilation (FMD%). In a scenario with no anticipated differences, FMD correlations were observed to be 0.8 at 12 weeks (95% CI: 0.72, 0.87). Therefore, assuming an auto-regressive structure, the correlation would be 0.64 at 26 weeks and 0.41 at 52 weeks. For a 25% relative difference (2% absolute difference) in FMD with 90% power and alpha 0.05, we would need just 30 subjects (comparing singing interventions to control). But, to compare the singing interventions to each other, using a delta of 0.015, we need 52 subjects for 90% power and alpha 0.05. To have 52 evaluable subjects (assuming a 25% dropout rate), we enrolled 65 subjects. With a change of 0.08 with 0.1 standard deviation in RHI, a sample size of 52 would yield a power of 81% with alpha 0.05. Therefore, in a cross-over design, we would be able to detect a change of 0.08 with power greater than 81% or we would be able to detect changes less than 0.08 with power equal to 81%. Similar remarks can be made for the Framingham RHI.

### Statistical Analysis

Statistical methodology included outlier detection and removal with estimation of standard models for a three-treatment, cross-over design. Outcome results for RHI, fRHI, brachial artery FMD, and HRV are displayed as summary statistics by type of intervention: control, video instruction, and coach (by a music therapist). Outliers exceeding 2.5 standard deviations from the mean were removed to avoid unduly influential results; generally, the number of outliers removed were few for each outcome. General linear models for a cross-over design with three treatments^28^ were fit for both absolute and relative differences of each outcome. Primary analysis models estimated effects for the subject, visit number, treatment, and carry-over, if any. Due to the COVID-19 pandemic, 20 out of 64 coaching intervention visits were conducted by the singing coach remotely. Therefore, secondary analysis models included the primary effects along with virtual coaching while the subject effect was partitioned by sex, race and age. Exploratory analyses included the secondary effects along with the video treatment partitioned by the four possible song choices (*Hey Jude, Jolene, This Land is Your Land, Amazing Grace).* These models were compared by rank-normalization (to assess the normality of the errors) and, similar results were found, i.e., the errors largely appeared to be approximately normal.

## Results

### Demographics

Sixty-five subjects were enrolled between 01/07/20 and 08/18/23. A total of 481 patients were assessed for eligibility, **Figure 1**. Eighty-two patients were excluded because they either did not meet the inclusion criteria or had at least one exclusion criterion for the study. Another 334 patients either declined to participate (n=204) or could not be reached after at least one attempt (n=130). One participant withdrew after the first visit and, therefore, was not included in the final analysis. Table 1, stratified by gender, summarizes the overall demographic characteristics of the 65 subjects enrolled. The mean age of participants was 67.7 (±0.8) years with 40% women. Most of the participants were non-Hispanic (98.5%) and identified as White (86.2%). The mean body mass index (BMI) of the participants was 30.0 (±1.0) with 49.0% categorized as obese and an additional 26.2% as overweight. Physical limitation was ascertained by self-report with 53.8% reporting some level of orthopedic limitation.

**Figure 1.**
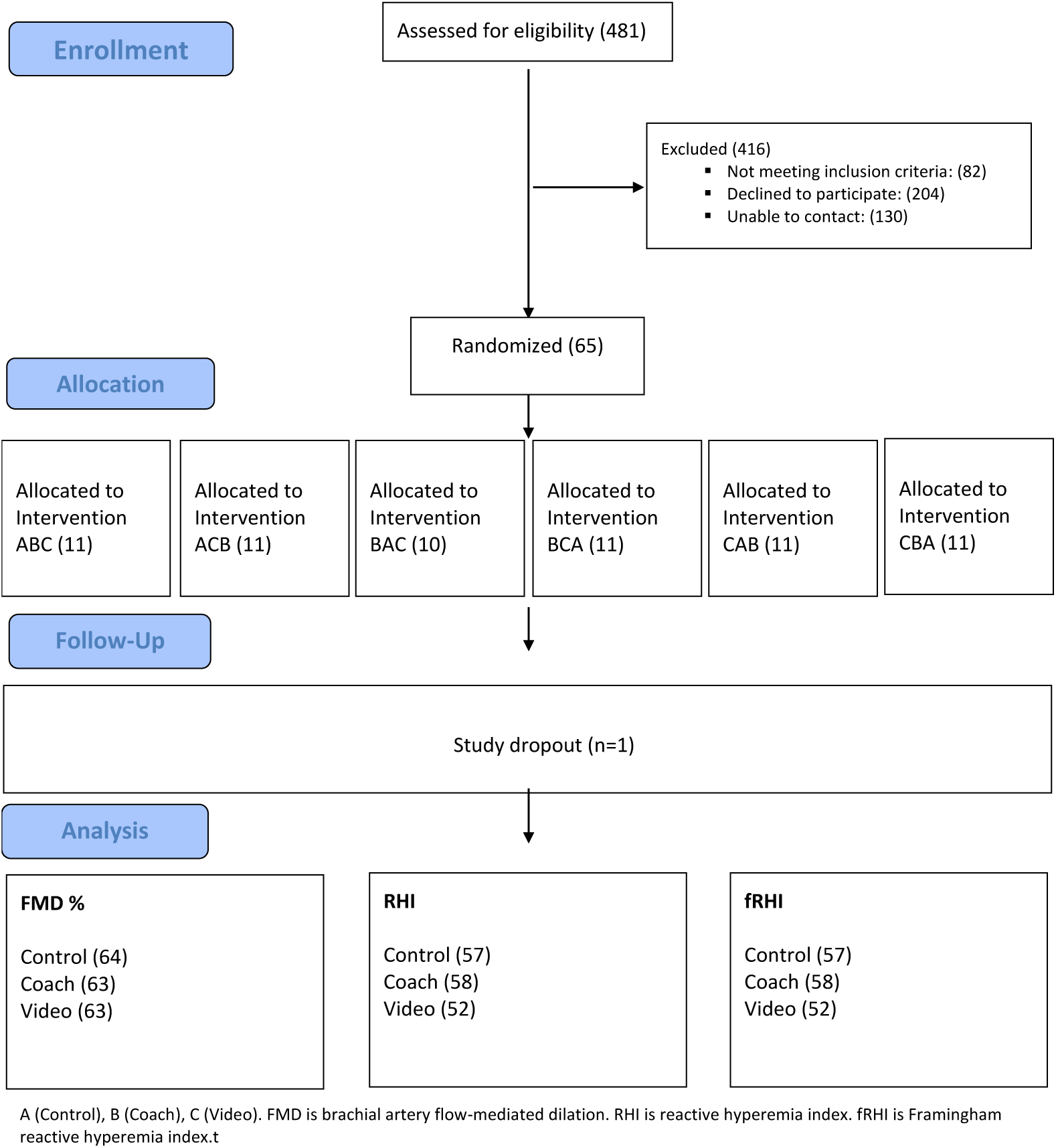
Consort diagram for randomized, cross-over, control trial.

**Table 1.** Baseline characteristics (n = 65)

| Table 1. Baseline characteristics (n = 65) |  |  |  |
| --- | --- | --- | --- |
|  | Total<br>n=65(col%) | Women<br>n=26(col%) | Men<br>n=39(col%) |
| **Age, mean ± SD, years** | 67.7 ± 0.8 | 68.3 ± 1.4 | 67.3 ± 1.0 |
| **Non-Hispanic ethnicity** | 64 (98.5) | 26 (100.0) | 38 (100.0) |
| **Race** |  |  |  |
| Black | 7 (10.8) | 3 (11.5) | 4 (10.3) |
| White | 56 (86.2) | 23 (88.5) | 33 (84.6) |
| Asian | 1 (1.5) | 0 (0.0) | 1 (2.6) |
| Unknown | 1 (1.5) | 0 (0.0) | 1 (2.6) |
| **History of coronary artery disease** |  |  |  |
| Myocardial Infarction | 41 (63.1) | 11 (42.3) | 30 (76.9) |
| Stent Placed | 47 (74.6) | 17 (70.8) | 30 (76.9) |
| Coronary Artery Bypass | 18 (27.7) | 5 (19.2) | 13 (33.3) |
| **Diabetes Mellitus** | 19 (29.2) | 9 (34.6) | 10 (25.6) |
| **Hypertension** | 49 (75.4) | 20 (76.9) | 29 (74.4) |
| **High Cholesterol** | 55 (84.6) | 21 (80.8) | 34 (87.2) |
| **Chronic Kidney Disease** | 10 (15.4) | 4 (15.4) | 6 (15.4) |
| **Chronic Respiratory Disease** | 18 (27.7) | 10 (38.5) | 8 (20.5) |
| **Heart Failure** | 12 (18.5) | 8 (30.8) | 4 (10.3) |
| **Prior Smoking** | 26 (40.0) | 12 (46.2) | 14 (35.9) |
| **BMI, mean ± SE** | 30.0 ± 1.0 | 30.5 ± 2.0 | 29.7 ± 1.1 |
| **BMI Category** |  |  |  |
| Underweight <18.5 | 4 (6.2) | 3 (11.5) | 1 (2.6) |
| Healthy weight 18.5 - 24.9 | 12 (18.5) | 5 (19.2) | 7 (17.9) |
| Overweight 25 to 29.9 | 17 (26.2) | 6 (23.1) | 11 (28.2) |
| Obese 30 or greater | 32 (49.2) | 12 (46.2) | 20 (51.3) |
| **Physical or orthopedic limitations** | 35 (53.8) | 15 (57.7) | 20 (51.3) |
| **Level of limitation (N=35)** |  |  |  |
| Minimal | 21 (60.0) | 7 (46.7) | 14 (70.0) |
| Somewhat | 12 (34.3) | 8 (53.3) | 4 (20.0) |
| Very | 2 (5.7) | 0 (0.0) | 2 (10.0) |
| **Ambulation aids (N=64)** | 6 (9.4) | 4 (15.4) | 2 (5.3) |
| **Assistive device use (N=35)** | 5 (14.3) | 4 (26.7) | 1 (5.0) |

All participants had coronary artery disease, one of the mandatory inclusion criteria. History of myocardial infarction was present in 63.1% of participants, coronary stenting in 74.6% and coronary artery bypass grafting in 27.1%. Amongst the comorbidities, dyslipidemia was most common and present in 84.6% of patients, followed by hypertension in 75.4%, and diabetes mellitus in another 29.2%. Forty percent of subjects were prior tobacco smokers with a mean cigarette pack year history of 15.5 (SE ± 2.6). All participants were non-smokers for at least one year prior to study enrollment.

Most characteristics including age, race, and comorbidities were balanced across both genders except for a few differences. More men had myocardial infarction (76.9%) and history of CABG (33.3%) compared to women (42.3% and 19.2% respectively). Diabetes was more common among women compared to men (34.6% vs 25.6%), as was heart failure (30.8% vs 10.3%). Included women had a more extensive tobacco use history (mean pack years of 19.8 ± 4.9) compared to men (11.8 ± 2.2).

### Vascular Function

The average pre intervention FMD% of the study participants was 4.2% (SE 0.19) highlighting the impaired baseline macrovascular endothelial function of the study population.^2^ Unbalanced Analysis of Covariance (regression) was used to calculate the crossover estimates between the two different singing interventions and control as absolute change in post and pre-intervention FMD%, Table 2. For absolute change in FMD%, the coaching intervention showed an estimated difference of −0.05 (SE 0.42, p=0.913) and the video intervention showed an estimated difference of −0.07 (SE 0.42, p=0.864) compared to the control intervention.

**Table 2.** Vascular function outcomes for singing interventions compared to control.

|  |  |  |
| --- | --- | --- |
| Parameter [Unit] | Absolute (post-pre) |  |
|  | Estimate (SE) | P-value |
| **fRHI** |  |  |
| Coach | 0.11 (0.18) | 0.570 |
| Video | 0.54 (0.19) | **0.005** |
| **RHI** |  |  |
| Coach | 0.09 (0.12) | 0.462 |
| Video | 0.13 (0.13) | 0.290 |
| **FMD [%]** |  |  |
| Coach | -0.05 (0.42) | 0.913 |
| Video | -0.07 (0.42) | 0.864 |
| Adjusted for the order of the intervention, carry over (if any), and period. |  |  |

Twenty-five (out of 192 or 13%) PAT measurements were not included in the final analysis due to equipment malfunction, excessive signal artifact, leaking tubing, or incomplete occlusion. Microvascular endothelial function, as assessed by RHI and fRHI, showed a statistically significant improvement for the video intervention only. Compared to control, the video intervention showed an estimated increase of 0.54 (SE 0.19, p=0.005) for absolute change in fRHI (30.9±11.1%, p=0.007, relative change; **Figure 2**). However, the estimated difference between the coaching intervention and control was not significant with an estimated increase of 0.11 (SE 0.18, p=0.570) for absolute change in fRHI. There was a statistically significant carry over effect observed with the video intervention (Estimate 0.69, SE 0.25, p=0.007) but not with the coaching intervention (Estimate 0.17, SE 0.25, p=0.503). There was no significant change in RHI with either singing intervention, Table 2. For absolute change in RHI, the estimated difference was 0.09 (SE 0.12, p=0.462) for coaching and 0.13 (SE 0.13, p=0.290) for the video intervention. Results of secondary and exploratory analysis models can be found in Supplementary Tables 2-4.

**Figure 2:**
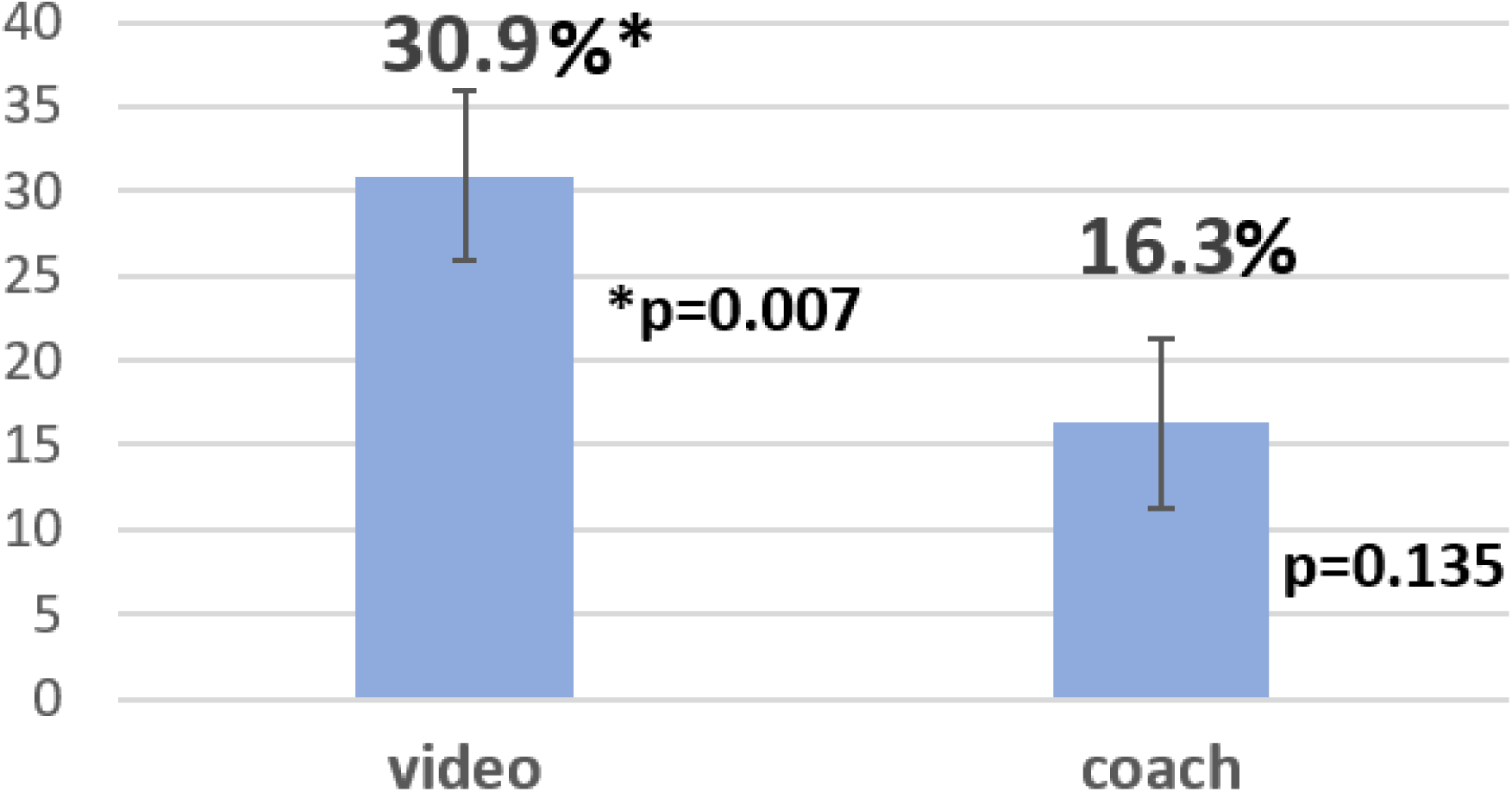
fRHI Relative Change (%) v. control (n=65)

### Heart Rate Variability

Heart rate variability (HRV) was assessed prior to, during, and after the intervention, with both during and post intervention values compared to pre intervention. Poor quality HRV recordings (5.7% pre-, 27.1% while singing, 21.2% post-singing) were excluded from the final analysis. The results for HRV are summarized in Tables 3 and 4. There were no significant changes in the estimated differences for the time domain HRV parameters, SDNN and RMSSD, for either singing intervention compared to control in either post- to pre- or during to pre-intervention comparisons.

**Table 3.**
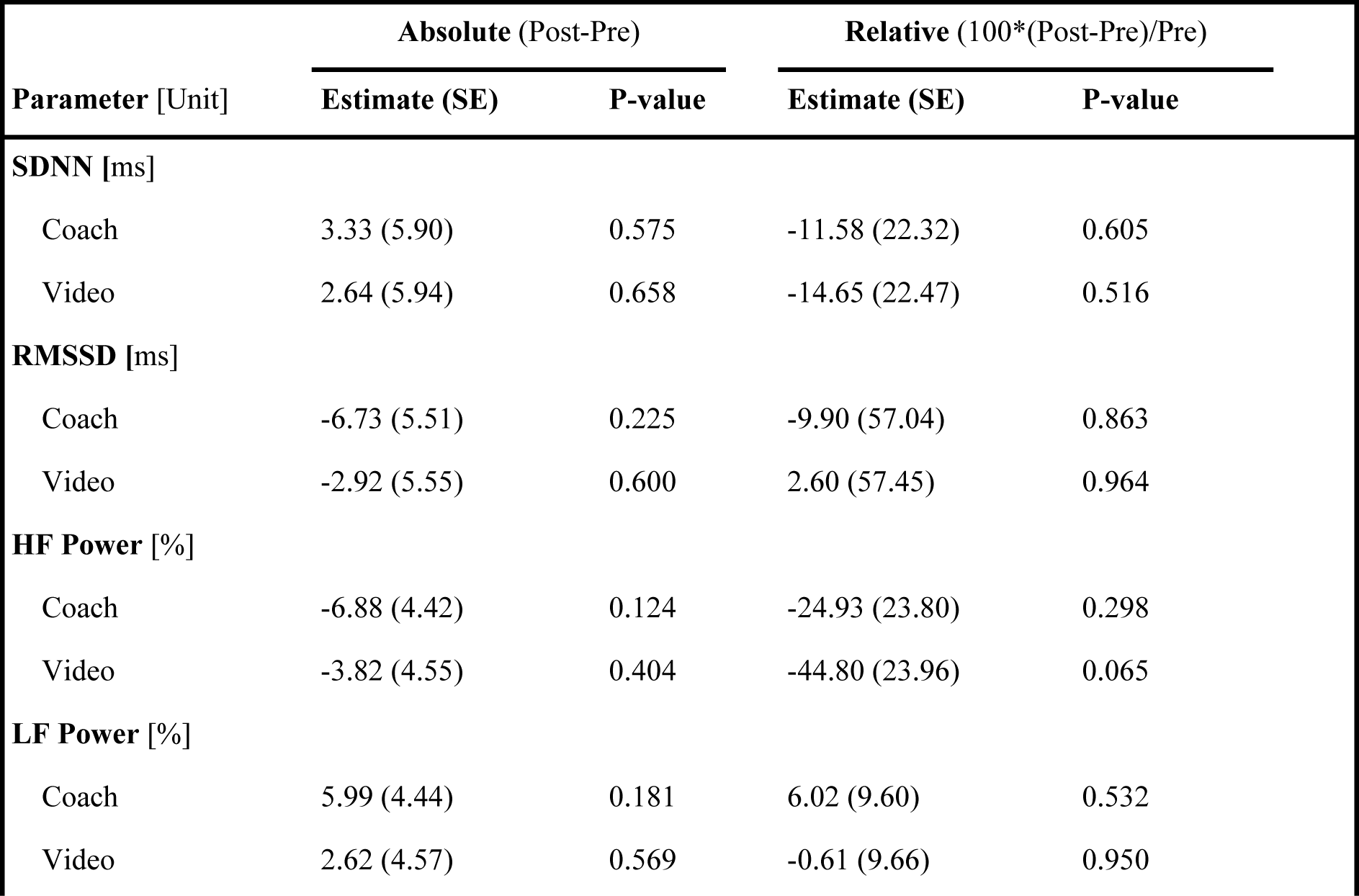
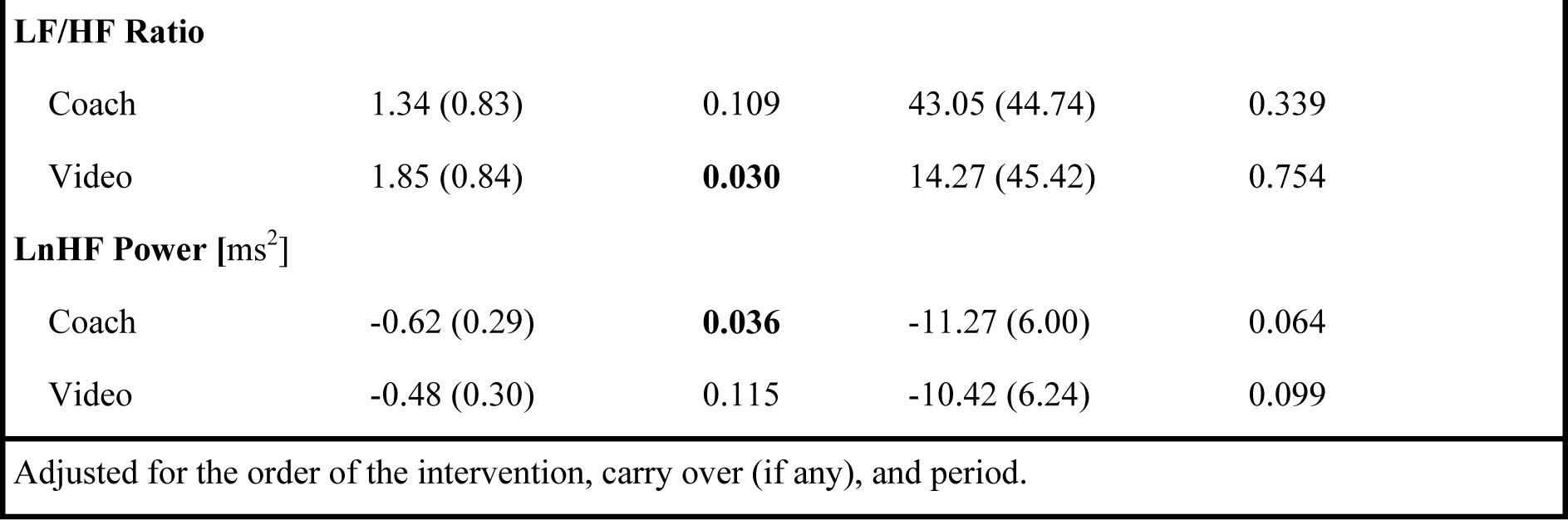
Heart Rate Variability (Post-Pre) outcomes for singing interventions compared to control.

**Table 4.** Heart Rate Variability (During-Pre) outcomes for singing interventions compared to control.

|  |  |  |  |  |
| --- | --- | --- | --- | --- |
| **Parameter [Unit]** | **Absolute (During-Pre)** |  | **Relative (100\*(During-Pre)/Pre)** |  |
|  | **Estimate (SE)** | **P-value** | **Estimate (SE)** | **P-value** |
| **SDNN [ms]** |  |  |  |  |
| Coach | -7.39 (5.93) | 0.217 | -32.17 (18.10) | 0.079 |
| Video | -2.56 (6.15) | 0.678 | -24.53 (18.78) | 0.195 |
| **RMSSD [ms]** |  |  |  |  |
| Coach | -7.72 (6.05) | 0.206 | 14.23 (50.31) | 0.778 |
| Video | -5.17 (6.16) | 0.403 | -14.47 (51.18) | 0.778 |
| **HF Power [%]** |  |  |  |  |
| Coach | 3.04 (3.81) | 0.427 | 10.48 (15.24) | 0.494 |
| Video | -5.59 (3.81) | 0.147 | -17.61 (15.25) | 0.252 |
| **LF Power [%]** |  |  |  |  |
| Coach | -4.21 (3.80) | 0.272 | -11.79 (8.95) | 0.192 |
| Video | 4.53 (3.87) | 0.246 | 1.94 (9.11) | 0.832 |
| **LF/HF Ratio** |  |  |  |  |
| Coach | 0.43 (1.00) | 0.670 | 1.96 (43.27) | 0.964 |
| Video | 2.80 (1.03) | **0.008** | 96.95 (44.53) | **0.033** |
| **LnHF Power [ms<sup>2</sup>]** |  |  |  |  |
| Coach | -0.55 (0.28) | 0.050 | -10.57 (5.52) | 0.060 |
| Video | -0.90 (0.29) | **0.003** | -15.63 (5.75) | **0.008** |
| Adjusted for the order of the intervention, carry over (if any) and period. |  |  |  |  |

For the frequency domain HRV parameters, the LF/HF ratio increased by 2.80 (SE 1.03, p=0.008), representing a relative change of 96.95% (SE 44.53%, p=0.033), for the video intervention compared to control (during to pre- change). When comparing post- to pre-intervention, the change in the LF/HF ratio remained significant (Estimate 1.87, SE 0.89, p=0.039). There was no significant change in LF/HF ratio with the coaching intervention. There were no significant changes in HF power and LF power for either singing intervention.

The estimated difference in natural logarithm HF (LnHF) power attained statistical significance for the video intervention (absolute change −0.90 ms^2^, SE 0.29, p=0.003; relative change −15.63%, SE 5.75, p=0.008) but not for the coaching intervention (absolute change −0.55 ms^2^, SE 0.28, p=0.050; relative change −10.57, SE 5.52, p=0.060) compared to control (during to pre-intervention comparison). However, when assessing post- to pre- change, it was the coaching intervention that showed a statistically significant difference of −0.62 ms^2^ (SE 0.29, p=0.036) in LnHF power compared to control. Results of secondary and exploratory analysis models for all HRV values can be found in Supplementary Tables 5-16.

Compared to the video intervention, the coach intervention led to higher BORG RPE scores (9.98±0.28 and 10.66±0.29, respectively) with a difference of 0.68 (SE 0.28), p=0.0167. Self-reported level of enjoyment (scale 1-10 with 10 most enjoyable) was higher for the coach intervention compared to the video intervention (9.3±0.13 and 8.5±0.21; p=0.0004).

## Discussion

In this randomized, single-blind, control, crossover study of singing interventions in older adults with established CAD, 30 minutes of singing along to a pre-recorded instructional video improved microvascular, but not macrovascular, endothelial function. To our knowledge, this is the first rigorously conducted trial of singing interventions to demonstrate acute improvements in microvascular endothelial function in a population with baseline endothelial dysfunction (mean FMD 4.2±0.33%). Findings advance the field of music medicine by providing insight into the mechanisms by which singing may improve health.

A hallmark of vascular aging is dysfunction of endothelial cells – the single layer of cells that lines all blood vessels and the heart and regulates exchanges between the blood and tissues. Healthy endothelial cells release nitric oxide which has vasodilatory and anti-thrombotic actions blocking the process of atherosclerosis. Endothelial dysfunction is considered an early precursor and common pathological feature of vascular diseases.^29^ For these reasons, direct measurement of vascular endothelial function is a powerful tool in translational science. These assessments have increasingly been applied in physiological studies to examine the mechanisms that underlie the acute or chronic impact of exposures that alter vascular function and risk for atherosclerosis (e.g., exercise training, smoking, hypercholesterolemia, hypertension, diet, medications). Vascular function strongly predicts cardiovascular events in patients with or without CVD,^9^ and endothelial dysfunction can be reversed by interventions known to reduce CVD risk.^10,11^ For example, there is an 8-13% lower risk of CVD events per 1% increase in BA-FMD. For microvascular function, the reactive hyperemia index (RHI) is provided by PAT. Meta-analyses show a 21% reduction in CVD events for every 0.1 natural log RHI (lnRHI) increase.^30^

The microcirculation, constituted by pre-arterioles, arterioles, capillaries, and venules, is responsible for most of the resistance to flow that modulates blood pressure and tissue perfusion and has been increasingly recognized as a key feature of CVD (compared to the larger, “macro” vasculature).^31^ Bonetti et al. measured digital pulse volume changes during reactive hyperemia in 94 patients without obstructive coronary artery disease and either normal (n=39) or abnormal (n=55) coronary microvascular endothelial function.^32^ Multivariable analysis identified coronary blood flow responses to acetylcholine as the only independent predictor of RHI.^32^ Therefore, despite being measured at the periphery, the RHI could be used as a potential non-invasive tool to identify patients with coronary microvascular endothelial dysfunction. Based on a systematic review and meta-analysis evaluating the prognostic magnitude of non-invasive vascular function testing, the estimated increase in fRHI of 0.54 (SE 0.19, p=0.005) in our singing video intervention arm translates into an impressive 40% reduction in CVD risk.^30^ While certainly promising, the next important step is to determine if this improvement in vascular function is sustained with a longer intervention period (weeks to months of singing).

There was a lack of improvement in large-vessel brachial artery FMD. It may be that the singing stimulus was not large (or intense) enough to effect change. Achieved mean Borg RPE score was 9.98 (±0.28) in the video intervention and 10.66 (±0.29) in the singing intervention (range for both was 6 to 16). These mean scores equate to light-intensity physical activity (less than 3 metabolic equivalents or METs or 25-44% VO2max).^26^ A meta-analysis concluded that higher intensity physical activities are likely needed to see a meaningful change in FMD in middle-aged and elderly people.^33^ In addition, more habitual activity over weeks to months, is more important from a prognosis standpoint and evaluation of sustained vascular adaptation should be tested in future clinical trials of singing. It is possible the timing of the FMD measurement after singing (or exercise) matters. For example, studies taking multiple FMD measurements after a bout of moderate or vigorous-intensity exercise show an immediate decrease in FMD followed by a normal or supranormal FMD response and ultimate normalization within 24-48 hours after exercise.^34^ Higher exercise intensities (>80% VO2max) resulted in a larger decrease in FMD post-exercise whereas most, but not all, studies of low-moderate intensity exercise reported an increase in FMD after exercise. We measured FMD approximately 40 minutes after singing. Based on extrapolation from studies of exercise, this would fall within the rise (of the initial drop) in FMD. Future studies of singing should explore this potential biphasic FMD response.

The lack of improvement in vascular function with the in-person coaching visit with a music therapist (compared to control) was unexpected, particularly since the intervention had a higher rating for enjoyment. It may be that a single visit singing with someone you just met could be anxiety-provoking, especially for amateur singers. A longer-term singing intervention or program could likely ameliorate this issue. Due to the COVID-19 pandemic, 20 out of 64 (31%) coaching intervention visits were conducted by the singing coach remotely. Therefore, secondary analysis models included the primary effects along with unanticipated virtual coaching. The virtual coaching sessions had a lower effect size (5.24%) compared to the in-person coaching sessions (14.2%) though we are underpowered to separate these for significance (Supplementary Table 1). Music structure (e.g., tempo, rhythm, dynamics) is complex, and it is possible that these elements could differentially impact physiological signals. With 40 song choices for the coaching visit, it is impossible to evaluate this. However, the video visit included just 4 song choices (and subjects selected 2). In a strictly exploratory analysis, we found that *Amazing Grace* had the largest effect size at 22.3% and *This Land is Your Land* had the lowest at 10.1% (Supplementary Table 1). These were not statistically significant findings but might be hypothesis-generating for future research in music medicine.

There was a carryover effect with the instructional video visit that is included in the analysis of crossover studies when detected. This likely means that a minimum of 2 days for potential washout from the previous visit was not long enough. One possible explanation for this carryover effect is the “earworm” – the experience of a song that repeats persistently in the mind – is a ubiquitous yet mysterious cognitive phenomenon, manifested as “inner singing” and may result in continued singing after the intervention.^35^ We did not ask subjects if they continued to sing in the days following their interventions, but this may be important to consider in future studies. Interestingly, functional MRI imaging of the brain in soprano Reneé Fleming in 2017 demonstrated that thinking about singing activated the brain to a larger extent than singing itself. This provides some proof of concept on the potential for earworms to have a physiological effect; however, more definitive studies are needed.

The acute HRV changes observed with the video intervention in our study, specifically increases in the LF/HF ratio and reductions in the LnHF power, are similar to those of exercise activity whereby exercise elicits an increase in sympathetic nervous system activity and parasympathetic withdrawal.^36^ In general, an increase in LF power implies a more dominant activity of the sympathetic nervous system (SNS) while an increased power in the high frequency band indicates stronger influence of the parasympathetic nervous system (PNS). Pagani et al. proposed the LF/HF ratio as an index of sympathovagal balance between the two nervous systems.^37^ Vagal activity is the major contributor to the HF component, and LnHF power is posited to reflect vagal tone,^38^ that decreased during our singing interventions. If we extrapolate what we know about regular exercise, it is likely that a longer intervention of singing (weeks to months) would lead to improved resting HRV due to enhanced autonomic nervous system balance.^39^ This may be especially important in an older population, as HRV is known to worsen with aging. These chronic alterations in HRV are likely more clinically meaningful than acute changes and should be incorporated into future studies of singing.

The strengths of the present study include the randomized cross-over with control intervention trial design. The cross-over design has the advantage of inherently controlling for subject-level variables (each subject serves as their own control). Furthermore, it did expose carry-over effects with the instructional singing video intervention. Because it is difficult to know what an appropriate washout period is for a singing intervention, future studies could consider alternative clinical trial designs and/or longer (than 7 days) of washout. Regarding HRV measurements, the accuracy of these measurements (with the equipment we used) during active singing, compared to rest, is questionable, possibly due to movement or perspiration.^40^ Therefore, commercial equipment used to assess HRV should satisfy industry standards in terms of signal-to-noise ratio and other performance metrics.^38^

## Conclusions

Singing along to a pre-recorded instructional video for 30 minutes improved microvascular, but not macrovascular, endothelial function, in older patients with known CAD. Singing should be considered as an accessible and safe therapeutic intervention in an older population who otherwise may have physical or orthopedic limitations hindering participation in traditional exercise. Future studies should explore the sustained vascular response to singing over weeks to months and explore the potential for “earworm” effects between visits.

## Data Availability

Additional data may be requested from the corresponding author after the initial peer-reviewed manuscript has been published.

## Disclosures

Research reported in this publication was supported by the National Center For Complementary & Integrative Health of the National Institutes of Health under Award Number R33AT010680. The content is solely the responsibility of the authors and does not necessarily represent the official views of the National Institutes of Health.

## Notes

### Competing Interest Statement

The authors have declared no competing interest.

### Clinical Trial

NCT04121741

### Author Declarations

Medical College of Wisconsin Institutional Review Board

